# Functional brain connectivity predictors of prospective substance use initiation and their environmental correlates

**DOI:** 10.1101/2024.05.29.24308134

**Authors:** Omid Kardan, Alexander Weigard, Lora Cope, Meghan Martz, Mike Angstadt, Katherine L. McCurry, Cleanthis Michael, Jillian Hardee, Luke W. Hyde, Chandra Sripada, Mary M. Heitzeg

## Abstract

**Background:** Early substance use initiation (SUI) places youth at substantially higher risk for later substance use disorders. Furthermore, adolescence is a critical period for the maturation of brain networks, the pace and magnitude of which are susceptible to environmental influences and may shape risk for SUI.

**Methods:** We examined whether patterns of functional brain connectivity during rest (rsFC), measured longitudinally in pre-and-early adolescence, can predict future SUI. In an independent sub-sample, we also tested whether these patterns are associated with key environmental factors, specifically neighborhood pollution and socioeconomic dimensions. We utilized data from the Adolescent Brain Cognitive Development (ABCD) Study^®^. SUI was defined as first-time use of at least one full dose of alcohol, nicotine, cannabis, or other drugs. We created a control group (*N* = 228) of participants without SUI who were matched with the SUI group (*N* = 233) on age, sex, race/ethnicity, and parental income and education.

**Results:** Multivariate analysis showed that whole-brain rsFC prior to SUI during 9-10 and 11-12 years of age successfully differentiated the prospective SUI and control groups. This rsFC signature was expressed more at older ages in both groups, suggesting a pattern of accelerated maturation in the SUI group in the years prior to SUI. In an independent sub-sample (*N* = 2,854) and adjusted for family socioeconomic factors, expression of this rsFC pattern was associated with higher pollution, but not neighborhood disadvantage.

**Conclusion:** Brain functional connectivity patterns in early adolescence that are linked to accelerated maturation and environmental exposures can predict future SUI in youth.

## Introduction

Substance use disorders are a major public health problem that affected more than 46 million individuals in the US in 2021 [1]. Early initiation of substance use during adolescence places youth at substantially higher risk for later substance use disorders [2, 3, 4, 5], as well as health risk behaviors [6], suicide [7], and mortality [8]. Adolescence is also a critical period in which brain networks go through notable maturational changes [9,10,11]. As such, the neural and psychosocial factors that can contribute to or predict substance use initiation (SUI) among teens have been the subject of multiple lines of inquiry in substance use research [e.g. 12,13,14]. However, network changes during this period are protracted and the maturation process is susceptible to influences from the environment surrounding the teen [15,16,17]. Patterns of acceleration or delay in brain development during adolescence have been found to be associated with negative environmental factors in both the socioeconomic and physical domains [18, 19]. Some of these same environmental factors are associated with early substance use [20,21]. It is therefore important to consider longitudinal brain changes and their environmental correlates when attempting to predict adolescent SUI. The current study evaluates whether longitudinal patterns in functional brain architecture in early adolescence can predict future SUI and how environmental disparities might play a role in these brain phenotypes [22].

The use of resting-state functional brain connectivity has grown rapidly within the past decade in clinical and developmental research [23, 24, 25], and substance use research has not been an exception [26, 27, 28]. The utility of the functional ‘connectome’, indexed as correlated functional magnetic resonance (fMRI) activity among all pairs of brain regions, comes from two important findings. The first is the observation that cognitive processes are better represented by coordinated activity across brain regions (i.e., networks) rather than activity in isolated brain regions [29]. The second is the discovery that resting-state fMRI connectivity (rsFC) patterns are highly individualized [30, 31] and the connectome can reliably predict individual differences in cognitive and psychiatric domains [32, 33]. Research in adolescents has since shown that functional brain connectivity patterns are robustly associated with individual differences in cognition and psychopathology [12, 34]. A study by Rapuano et al., 2020 [12], for example, showed that rsFC can predict a “risk-seeking” dimension in 9–10-year-olds enrolled in the Adolescent Brain Cognitive Development (ABCD) Study^®^.

In the current study, we investigated whether whole-brain rsFC at pre- and early-adolescence timepoints (ages 9-10 and 11-12, respectively) can be used to distinguish youth who later initiated use of alcohol, nicotine, cannabis, or other drugs from their peers who did not (Study 1). The use of two fMRI timepoints (two years apart) that precede the initiation of substance use enabled an examination of developmental shifts in rsFC (e.g. accelerated/ delayed patterns) that may precede substance use.

Associations between the neighborhood environment and measures of cognition and mental health have been reported in multiple studies[35, 36, 37, 38], but neighborhood factors have been generally understudied in neuroscientific research on substance use. Specifically, neighborhood socioeconomic (SES) factors, including area deprivation index (ADI, which captures the economic disadvantages and poverty-related statistics in the area [39]) and measures of neighborhood safety and crime have been tied to the structure and function of the human brain cross-sectionally and longitudinally [37, 40, 41, 42, 43] while also being linked to substance use [44, 45]. In addition, physical neighborhood variables capturing pollution, particularly neurotoxin-containing types of pollution such as fine particulate matter (PM2.5) and lead, have also been shown to be associated with functional and structural brain development [46, 47, 48, 49, 50] and shifts in neural plasticity and the potential timing of sensitive periods [51]. However, their relation with dimensions of SUI have not been well-studied. Understanding whether these factors are associated with brain phenotypes that are predictive of substance use can shed light on the contributions of the environment to development of substance use behaviors. Therefore, in an independent sub-sample, we explored correlations between the rsFC pattern identified in the first study and neighborhood pollution and socioeconomic factors.

In summary, the current study aims to find the patterns of functional brain connectivity that precede adolescent substance use, a strong risk factor for substance use disorders, and explore the environmental correlates of these brain signatures. We utilize multivariate methods to ask this question in a large and heterogeneous longitudinal sample. In doing so, we will inform research on the trajectory of substance disorders and their neurodevelopmental antecedents.

## Methods and Materials

### 1.1. Participants

We utilized data from the Adolescent Brain Cognitive Development (ABCD) Study^®^ [52, 53], which is an ongoing longitudinal study of 11,867 children across 21 sites in the US. Participants were enrolled in the study between 9–10 years of age, and the study involves MRI acquisitions every two years and substance use assessment surveys every year (complemented with brief mid-year surveys). The starting sample was 52.2% male and composed of 52.0% non-Hispanic White, 15.0% Black, 20.3% Hispanic, 2.1% Asian, and 10.5% Other participants.

Across our analyses, we utilized all available substance use survey measures from baseline (Y0) to Year 4 (*N* = 4,754 currently available for Y4). The final sample size for this study was 3,801 participants with suitable neuroimaging data at both baseline (ages 9–10) and Year 2 (ages 11–12) (see exclusion criteria in section 1.2). This sub-sample was 60.1% non-Hispanic White, 10.1% Black, 18.2% Hispanic, 1.5% Asian, and 9.9% Other, which underrepresents Black/African American participants compared to the overall sample. The sub-sample was comparable to the overall sample in terms of substance use initiation with 8.0% (N = 949) in the overall and 8.4% (N = 321) in the sub-sample, respectively. The sub-sample included fewer male participants compared to the overall sample but was balanced in terms of biological sex (50.4% male). Finally, the included sample was slightly higher on parental education (Mean = 17.4 years in the subset compared to 17.2 years in the overall sample; two-sample *t* = 4.67, *p* < .001) and also higher on household income (Mean of income bracket midpoints = $79,360 compared to $71,890 in the overall sample; two-sample *t* = 6.72, *p* < .001).

Since the goal of our analysis was to predict future SUI, participants who had already initiated substance use by Y2 (i.e., within the 9-12 years of age) were not included in the analysis (N = 88 within the sub-sample). The first study involved a total of 461 participants belonging to two groups. The first group comprised participants who had initiated use of any drugs or alcohol (only full doses were considered not single puff or sip) during the Y3-Y4 study periods (ages 12-14) but not earlier (*N* = 233; *SUI group*). The second group comprised participants who did not initiate substance use and were matched with the SUI group in terms of age, biological sex, race/ethnicity, household income, and parental education (*N* = 228; *control group*) [see **supplementary section 1** for the matching algorithm]. Neither group had initiated substance use by the time of their Y0 and Y2 fMRI scans and their only SUI-related difference was *later* use of substances vs. not.

The second study utilized the remainder of participants with suitable MRI data and non-missing family and neighborhood variables who were not included in the two groups used in the first study and had not initiated substance use at least by the end of the second neuroimaging timepoint (*N* = 2,854).

### 1.2. fMRI processing pipeline and exclusions

For both Y0 and Y2 neuroimaging sessions, resting-state fMRI was acquired in four separate runs (∼5 min per run, full details are described in [54]). Briefly, images were acquired at a spatial resolution of 2.4 mm isotropic and a temporal resolution of TR = 800 ms. The entire data pipeline was run through automated scripts on the University of Michigan’s high-performance cluster and is described in detail elsewhere [24]. Key features of the pipeline include FreeSurfer normalization, ICA-AROMA denoising, CompCor correction, and censoring of high motion frames with a 0.5 mm framewise displacement threshold. Visual quality control (QC) was conducted to assess registration and normalization steps. Participants with at least one resting-state run at both baseline and Y2 that passed QC and more than 250 degrees of freedom left in the BOLD timeseries after confound regression and censoring were included in the rsFC analysis. For the rsFC analysis, the fMRI data were spatially down-sampled to 333 parcels in the Gordon parcellation for the cortical regions 55] augmented with 54 subcortical parcels [56] and 31 cerebellar parcels [57]. Functional connectivity matrices were generated using these 418 parcels for each participant for baseline and Y2 data. To maximize data per participant, we used all available eligible runs for each participant at each timepoint. [Also see **supplementary section 4**].

### 1.3. Variables

#### Substance use variables

Variables from two ABCD Release 5.0 tables were used to indicate if and when a full dose of substance was used by each participant. The full list of these items can be seen in our shared analysis script at https://github.com/okardan/future_SUI_rsFC (102 items from the su_y_sui table and 26 items from the su_y_mypi table were used). Briefly, we used the timeline follow-back (TLFB) yearly survey and midyear mypi and xskipout survey items to determine the initiation of the use of any cannabis, nicotine, alcohol, or other drugs across different methods of administration. If the combined count of total days of any type of nicotine use (cigarette, e-cig, hookah, etc.), cannabis use (smoked/vaped), alcohol use (any drinks containing alcohol), or other drugs (e.g., synthetic, inhalant, tranquilizer, etc.) during the entire cumulative period between sessions was equal to or greater than 1, the participant was flagged as having used a substance during that year. Participants were counted towards SUI if they reported having used a full dose of nicotine, cannabis, alcohol, or other drugs in their brief mid-year survey even if they did not report it later in the yearly TLFB.

#### Environmental variables

In the second study, we explored the correlations between the rsFC pattern identified in the first study and the neighborhood variables that have been previously tied to substance use behaviors, adolescent neural development, or both. Neighborhood perceived safety was determined by averaging the perceived safety reported by the youth and parents (from tables ce_p_nsc and ce_y_nsc) of the ABCD Release 5.0. The perceived safety items were averaged over all 4 years within each participant but years with missing values were ignored in the calculation of the mean. Neighborhood crime rate was based on violent crimes and drug sales in table led_l_crime at baseline, which were compiled by the Inter-University Consortium for Political and Social Research from FBI data (census tract level). The fine particulate matter (PM2.5) and NO2 air pollution were based on led_l_pm25 and led_l_no2 tables at baseline, respectively (annual average concentration at 1×1 Km^2^ around the primary residence). The lead exposure variable was based on table led_l_particulat (annual average of Lead in ng/m3 at 50m of primary residence at baseline). The area deprivation index for each participant’s address was obtained from the weighted sum score in the table led_l_adi at baseline (derived from the American Community Survey at census tract resolution).

#### Family variables and covariates

The parental history of alcohol and drug use variables were parental reports from table mh_p_fhx indicating either parent with drug use or alcohol use problems. Family conflict was the average of the youth and parental report on the Conflict subscale of the Family Environment Scales reported in tables ce_y_fes and ce_p_fes. Covariates included data collection site and head motion in the scanner during the resting-state fMRI (mean frame displacement). Other covariates were from the abcd_p_demo table including parents reports on the participant’s biological sex, interview age, race/ethnicity, parental education (highest education), and household income. Family ID was not considered in the main analyses to maximize sample sizes, but supplementary analyses where only one sibling from each family was randomly retained yielded very similar results for both studies (see supplementary results section 2).

### 1.4. Statistical analyses

In study 1, we utilized a partial least squares (PLS) multivariate analysis [58, 59, 60, 61, 62, 63] to distinguish the SUI group from the matched Control group based on their resting state functional brain connectivity in the years prior to SUI. In brief, the analysis results in Latent Variables (LVs) that are linear combinations of functional connections across the whole brain whose combinations are differentially instantiated across groups (SUI and control) and years (baseline and Y2). [see **supplementary section 2** for details].

In study 2, we projected the identified rsFC pattern from study 1 onto the fMRI connectivity data in the remainder of participants (i.e., not among the SUI and Control groups) who also had not reported any substance use at baseline or Y2 and had complete data for the environmental and family variables (N = 2,854). [see **supplementary section 3** for details]. We then regressed these brain scores on the variables of interest (see section 1.3) in two ways. In both analyses, the model included random intercepts for the data collection site, as well as fixed-effects covariates age, sex, mean frame displacement, household income, and parental education. The difference between the two analyses was whether race/ethnicity was also included as covariates. Race/ethnicity variables are generally included to represent some of the systematic adversities of the marginalized minority groups in the US ([64], see field-wide debates about using race/ethnicity variables as proxies for exposure to unequal levels of adversity, discrimination, and opportunity among marginalized communities in the US [65]). However, because some of the neighborhood variables of interest in the current study including perceived safety, neurotoxins, and ADI may fall within those very adverse experiences, we included the version of the analysis where the variance explained by race/ethnicity was not accounted for.

## Results

### 2.1. Study 1: An rsFC acceleration-like pattern distinguishes prospective SUI from control

There were no significant differences between the SUI group and the control group in any of the matched categories (age: *p* = .681; sex: *p* = .988; income: *p* = .473; Education: *p* = .888; non-Hispanic White: *p* = .508; Black: *p* = 0.849; Hispanic: *p* = .812). Additionally, there was no difference between the two groups in terms of their mean head motion during scans (frame displacement: *p* = .471).

We utilized PLS regression to find the rsFC pattern across two timepoints (9–-10 and 11–-12 years of age) that reliably distinguished prospective SUI teens (N = 233) from matched controls (N = 228). The PLS analysis resulted in a significant primary latent variable (LV1: *p* < .001, R^2^ = .139, σ_XY_ = .515). Subsequent LVs were not statistically significant based on the permutation tests and are not discussed further. This primary LV is shown in the two panels of Figure 1. There is a pattern of connectivity that is expressed less in the control group (green bars) compared to the SUI group (yellow bar) during both 9–10 years of age and 11–12 years of age. The rsFC pattern is relatively wide-spread, but most strongly expressed as higher connectivity within the cingulo-parietal and between the two components of the somato-motor network, and lower connectivity within the cingulo-opercular network, between the cingulo-opercular network and the subcortical and the auditory networks, between auditory and subcortical networks, and between the cingulo-parietal and the default mode networks.

**Figure 1.**
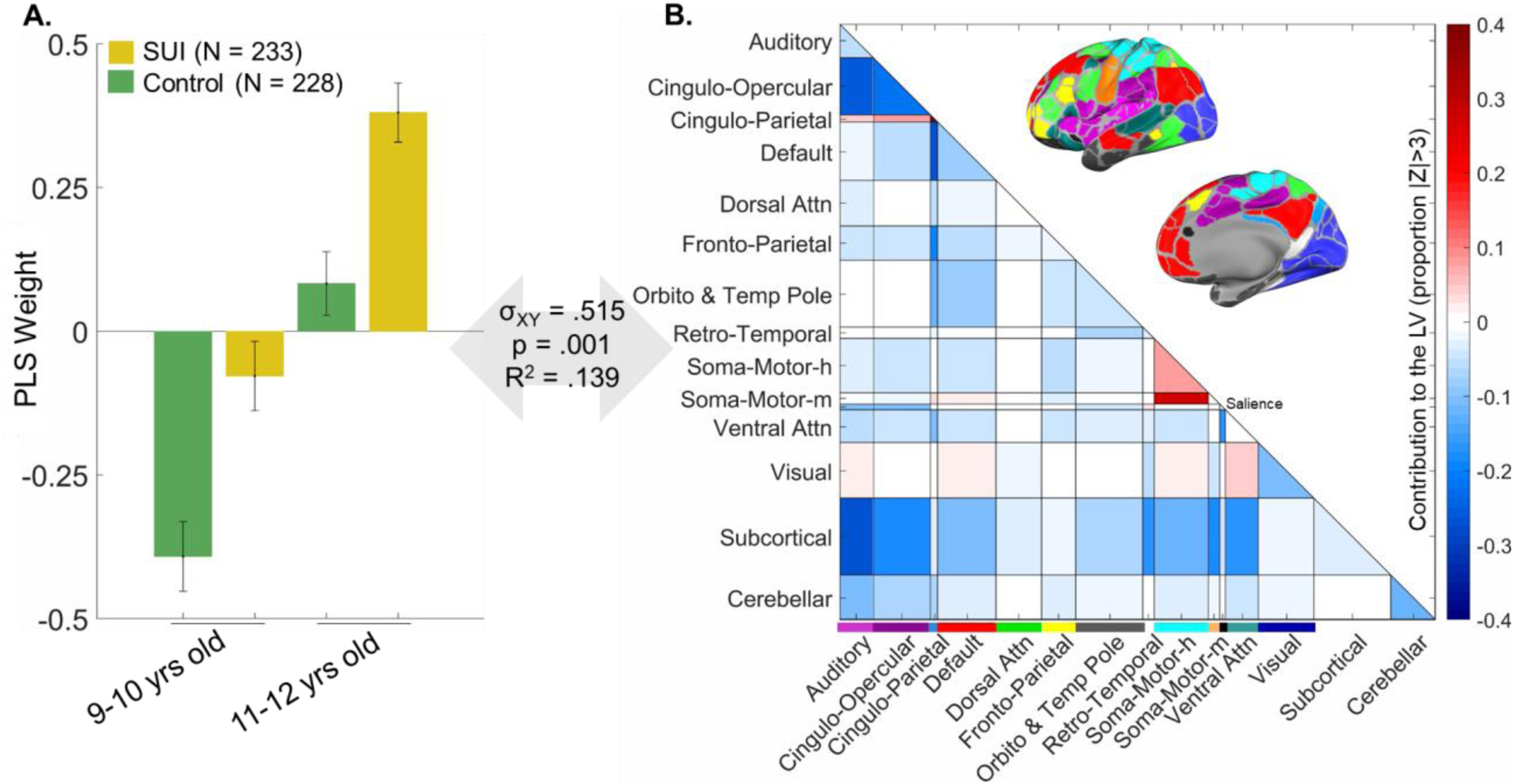
First latent variable (LV) from the partial least squares (PLS) analysis. In panel A, the group by time PLS weights in the latent variable are shown. The SUI group is in yellow, and the control group is in green; the first two columns show the loadings at 9–10 years of age and the second pair of columns show the loadings at 11-12 years of age. Panel B shows the extent to which the connections across and within brain networks load onto the PLS latent variable. The cortical networks are color-coded, and their anatomical mapping can be seen in the brain surface maps. Networks are defined according to [55]; the ‘unassigned’ set of connections in [55] is labeled here as ‘Orbito and Temp Pole’ as it falls in the orbitofrontal cortex and the temporal pole. The somato-motor networks are labeled with ‘h’ for hand and ‘m’ for mouth. The cross-block covariance is shown as σ_XY,_ and represents the squared singular value for this LV divided by the sum of squared singular values for all LVs. P value is determined by 1000 permutations, and the R^2^ here is the squared Pearson’s correlation (between the left and right-hand sides of the LV (i.e. U and V scores). Z values in the right panel and the error bars in the left panel are estimated using 1000 bootstraps (see supplementary methods).

Notably, the pattern shows an increased expression for both the SUI and control groups from younger to older ages. Given the fact that the groups are matched for age (i.e., no age difference between the SUI and control groups; t = .411, *p* = .681), this emerged age-dependent contrast between the two groups in the PLS latent variable should be investigated. To further substantiate this temporal component, we conducted a post-hoc PLS analysis where the mean rsFC across the two timepoints was removed. In this way, the longitudinal aspect of the data is removed and instead the rsFC at the two timepoints are averaged for each participant (i.e., as if these were measured across two same-day fMRI scan sessions). Therefore, finding a significant PLS LV distinguishing the SUI and control groups would require a time-independent rsFC difference across the youth. Supporting the importance of the longitudinal information in rsFC for differentiating the SUI and control groups, no significant brain saliences emerged from this analysis (permutation *p* = 0.372 for the primary LV, N.S.).

Importantly, the rsFC pattern identified in Figure 1 distinguishes prospective substance users from their non-using peers, but in our definition of SUI we did not separate different types of substances. As such, it is not clear whether the pattern is equally predictive of different groups of substances or mainly driven by only one group. In order to assess the degree to which the rsFC pattern is transdiagnostic of any substances or more specific to one type, we plotted how discriminative the expression of the rsFC pattern is for different types of substances (alcohol, nicotine, cannabis, other drugs, and poly-substance) versus the control group. As can be seen in Figure 2, prospective users of cannabis, nicotine, and other drugs, as well as those who used substances from multiple of these categories (poly) have significantly higher expression of the pattern than the control group, thus indicating some degree of common brain phenotype for multiple types of substances. However, the discrimination of the rsFC pattern is not homogenous across substance types. Specifically, later use of only alcohol is not significantly distinct from the control group. Given that most participants in the SUI group belonged to nicotine-only or the poly-substance sub-groups, and the alcohol-only and cannabis-only users were limited (i.e., many of the youth who initiated cannabis or alcohol use also started using substances of other categories), these potential differences should be further investigated when larger sample sizes for each type of substance may be available.

**Figure 2.**
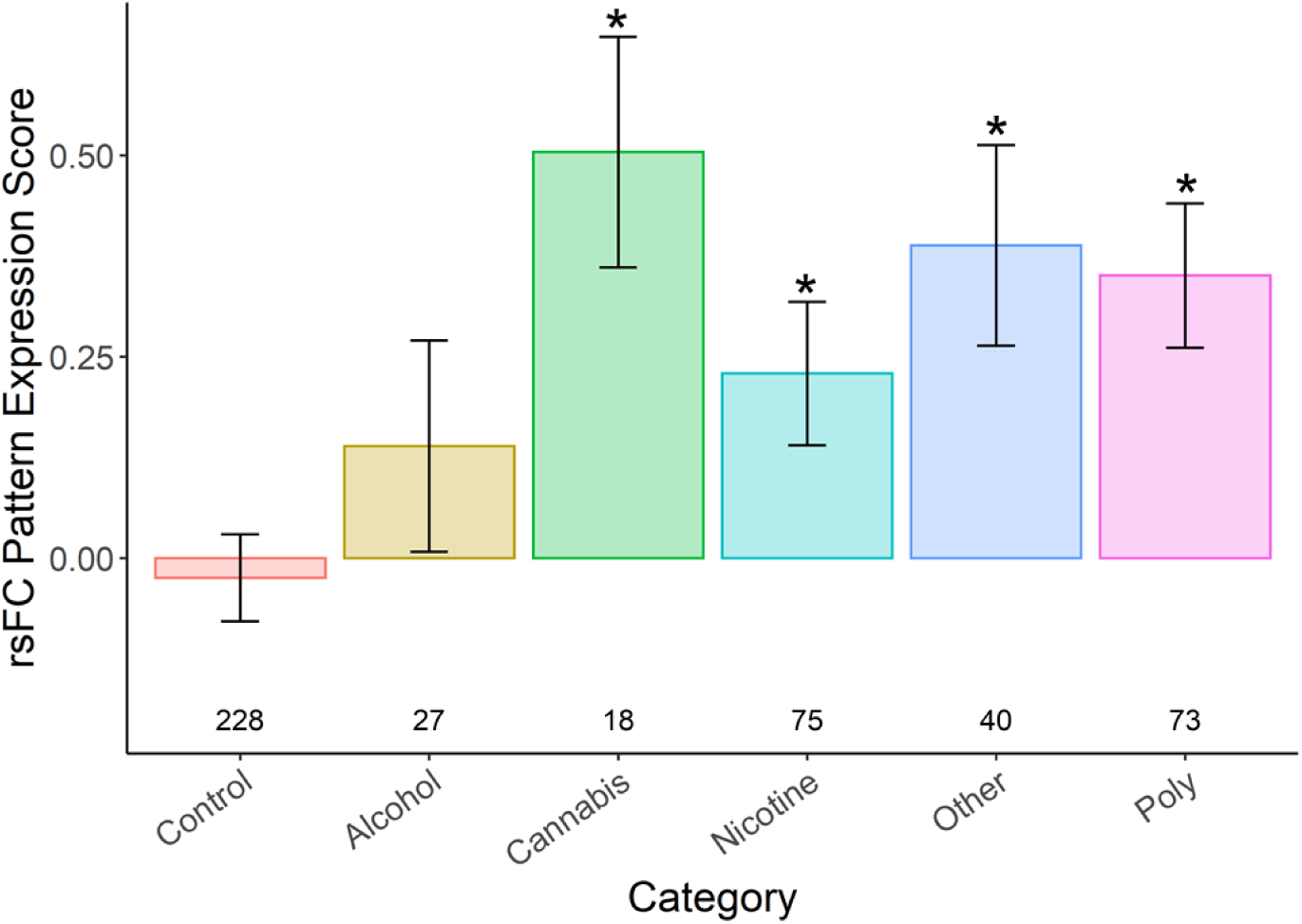
The expressed resting-state functional connectivity (rsFC) pattern across different categories of substances compared to the control group. The bars show mean expression score in the control group and each sub-group of the SUI group (see Methods section 1.4). * indicates *p* < .05 compared to the control group after correction for multiple pairwise comparisons. The number of participants in each (sub)group is shown below the bars.

### 2.2. Study 2: Environmental correlates of the rsFC pattern for prospective SUI

To better characterize this age-dependent rsFC pattern, we projected the pattern onto the fMRI connectivity data of the remainder of participants (who were not in either the SUI or the control group in the first study; N = 2,854). These participants had rsFC data with sufficient quality at both 9–10 and 11–12 years of age, and had not initiated substance use during this time, but may or may not have initiated use afterwards (undetermined because ABCD Release 5 is a partial data release).

Table 1 shows the associations between the projected rsFC-SUI pattern and family and environmental variables, adjusted for covariates (Model 1 does not include race/ethnicity as covariates, Model 2 does include race/ethnicity as covariates). In the family domain, we found that higher expression of this age-related rsFC pattern in the connectome may be associated with higher parental history of drug use (not significant after Holm-Bonferroni correction), but it was not significantly associated with parental history of alcohol use or family conflict.

**Table 1.** Results of regressions with the projected rsFC pattern as the outcome in the Study 2 youth. β is the standardized estimate for the regression coefficient. The difference between Model 1 and 2 is whether race/ethnicity are included as covariates or not. Note: N = 2,854 in both models. Grey highlight shows *p* < .05 within the isolated regression, and addition of * shows *p* < .05 after Holm-Bonferroni correction for all of the regressions. Hx = history.

| Predictor | Model 1<br>*Random effects: Site*<br>*Covariates: Age, Sex, Head motion, Family income, Parental Education* |  |  | Model 2<br>*Random effects: Site*<br>*Covariates: Age, Sex, Head motion, Family income, Parental Education, Race/Ethnicity* |  |  |
| --- | --- | --- | --- | --- | --- | --- |
| | $\beta$ | t | p | $\beta$ | t | p |
| **Family** |  |  |  |  |  |  |
| Parent Hx of Drug Use | 0.029 | 1.873 | 0.061 | 0.034 | 2.260 | 0.024 |
| Parent Hx of Alc Use | 0.015 | 0.960 | 0.337 | 0.020 | 1.309 | 0.191 |
| Family Conflict | 0.004 | 0.266 | 0.790 | 0.003 | 0.184 | 0.854 |
| **Neighborhood (Pollution)** |  |  |  |  |  |  |
| PM2.5 | 0.098 | 4.161 | <0.001* | 0.083 | 3.569 | <0.001* |
| NO2 | 0.049 | 2.106 | 0.035 | 0.012 | 0.542 | 0.588 |
| Lead | 0.046 | 2.126 | 0.034 | 0.012 | 0.572 | 0.567 |
| **Neighborhood (SES)** |  |  |  |  |  |  |
| Area Deprivation | 0.029 | 1.338 | 0.181 | -0.000 | -0.007 | 0.994 |
| Perceived Safety | -0.008 | -0.489 | 0.625 | 0.014 | 0.883 | 0.377 |
| Crime Rate | 0.039 | 1.133 | 0.257 | -0.006 | -0.191 | 0.848 |

In the neighborhood pollution domain, higher neighborhood air pollution, specifically fine particulate matter concentration (PM2.5), was significantly associated with stronger expression of the age-related rsFC pattern across participants. Other pollutants, NO2 and lead exposure, may have a modest relationship with the expression of the rsFC pattern but require further investigation (not significant after Holm-Bonferroni correction). In the neighborhood SES domain, we did not find a significant association between the rsFC pattern and area deprivation index (ADI), youth and parental reports of neighborhood safety, or the crime rate in the neighborhood in either regression model.

Finally, we conducted two sensitivity analyses which showed that the results are robust to the number of fMRI runs per participant and exclusion of family members [see **supplementary section 4**].

## Discussion

Early initiation of substance use during the teen years places youth at substantially higher risk for later substance use disorders and negative health outcomes. In the current study, we found an age-dependent pattern of functional brain connectivity across ages 9–10 and 11–12 years that differentiated prospective substance users from controls. The pattern of connectivity was expressed more in youth who will initiate substance use by age 14 compared to youth who will not, and more at older ages in both groups. Furthermore, given the age-related pattern in the PLS latent variable (Figure 1) and the fact that the groups are matched for age, prospective substance users look like they are shifted ahead in these rsFC patterns compared to their non-substance-using peers. The temporal pattern therefore may suggest an accelerated rsFC maturation in the youth a few years before they initiate substance use. We then assessed the neighborhood environmental correlates of this acceleration-like rsFC pattern in the remainder of the sample. Our results showed that this pattern was associated with dimensions of pollution, especially concentration of fine particulate matter in the air proximal to the participants’ residences. These findings are strengthened through the use of a substantial set of youth matched across demographic variables (N=461) with prospective data on which youth will go on to early substance use initiation, the use of a second subsample (N=2,854) to separately examine the environmental correlates of these brain patterns, and sensitivity checks to establish the robustness of the data, all within a large-scale, heterogenous sample of youth.

The current study is novel as few studies have examined associations between the whole-brain rsFC and adolescent SUI. However, some of the present results do fit with converging work across related research. Prior literature investigating rsFC in individuals with substance use disorders suggests that reduced connectivity within networks underlying cognitive control (e.g., fronto-parietal, cingulo-opercular, cingulo-parietal, and salience networks) is associated with precursors to substance use disorder (e.g., impulsivity), is found in substance withdrawal states, negatively predicts treatment outcome, and appears to be improved by substance use disorder interventions [66]. A recent meta-analysis of case-control studies similarly found that substance use disorder diagnosis was associated with reduced connectivity within and between the fronto-parietal and salience networks, as well as reduced connectivity between the limbic and default mode networks [67]. Several studies in relatively small samples have investigated rsFC correlates of substance use in youth. One study found that reduced connectivity between a network of regions involved in cognitive control and a network of subcortical regions was associated with problematic substance use in adolescents [68]. Other small studies have linked earlier SUI to greater connectivity between the fronto-parietal network and the limbic network [69] and greater connectivity between the fronto-parietal netwrok and nucleus accumbens specifically [70]. Finally, a study in the large IMAGEN consortium sample identified patterns of connectivity that predicted problematic alcohol use, primarily in females [71]. Although connections that positively and negatively predicted alcohol use were widespread across many networks, the somato-motor, salience, and subcortical networks were most prominent. Our study extends this literature and suggests that altered patterns of functional connectivity are not only related to the pathophysiology and maintenance, but also the prospective early initiation, of substance use.

Our finding that SUI was generally negatively associated with connectivity within the cingulo-opercular network, which is linked to cognitive control is consistent with findings on rsFC in substance use disorders [66, 67]. However, the higher connectivity within the cingulo-parietal network does not fit this interpretation. Additionally, given that the pattern of connectivity we identified appears to increase with age, our findings are somewhat at odds with the typical interpretation of reduced connectivity in these networks as reflecting reduced cognitive control [66]. Cognitive control shows substantial improvements with age throughout adolescence [72]. Therefore, the SUI-linked pattern of negative connectivity we identified is unlikely to represent a signature that indicates poorer cognitive control, as it displays the opposite developmental pattern, and may instead reflect a distinct signature of accelerated maturation in these networks that is triggered by environmental exposures. Our findings are also consistent with findings highlighting the importance of somato-motor and subcortical networks for predicting alcohol problems in adolescence [71] and the prior finding [68] that reduced connectivity between cognitive control and subcortical networks is associated with adolescent substance use. While our finding that SUI was strongly predicted by connectivity of the somato-motor network may initially seem surprising, it accords with recent data emphasizing that this network is particularly sensitive to variation in household socioeconomic resources in the ABCD Study [73] and is implicated in impulsivity, cognitive functioning, and transdiagnostic psychopathology in adults [74]. Interactions between cognitive control and subcortical systems are also critical for salience processing and emotion regulation, sensitive to environmental risk factors of SUI, and predictive of mental health across the lifespan [75, 76, 77].

Research on environmental influences on the pace of brain development has shown that experiences of threat and potentially also deprivation could accelerate the brain maturation processes [78, 18], despite debates about the boundary conditions under which stress and disadvantage may accelerate or delay neurodevelopment [79, 19]. Some of these accelerated development findings are then interpreted as being adaptive from the perspective of the life-history theory. For example, accelerated brain development in contexts of threat, disadvantage, and unpredictability may allow youth to navigate their environment and regulate their emotions more independently at an earlier age [80]. However, this acceleration may shorten periods of peak neural plasticity, limiting subsequent learning and adaptation to changing contexts and incurring long-term costs to health and behavior [but see 65]. The current study suggests factors contributing to the accelerated maturation of the functional brain connectivity in early adolescence may also contribute to later substance use initiation. Additionally, our findings suggest that research in this domain could benefit from expanding to include the neighborhood physical environment as a potential dimension of adversity or deprivation (or added as an exposure factor), with implications for adolescent substance use. This finding also continues to identify structural, rather than only familial, targets for prevention and intervention.

A possible limitation of the present study is that substance use data was self-reported, which may limit the accuracy of actual use behavior. Stigma around substance use or concerns about negative consequences of disclosing substance use may contribute to hesitancy in endorsing use. Other large-scale, national studies, including Monitoring the Future and the Youth Risk Behavior Surveillance System surveys, have used self-report substance use behavior. Converging findings across these studies suggests accuracy in self-reported substance use. However, a recent study using both self-report substance use data and toxicological hair assessment in 696 participants from the ABCD Study sample found that 10% of these participants had hair toxicology results inconsistent with self-reported substance use [82]. Thus, results from this work suggest that substance use may be underestimated from self-report assessments. Another limitation is that use across the different categories of substances was unequal and thus did not allow for full understanding of how the expression of rsFC patterns is different for types of substances (alcohol, nicotine, cannabis, other drugs, and poly-substance) versus the control group. The underrepresented Black/African American youth in the final sample in both study 1’s groups, as well as in the remainder of the sample used for study 2 was another limitation. As stated elsewhere [83, 84], maximizing the large sample size of the ABCD Study while simultaneously ensuring data quality and representativeness remains a challenge.

In conclusion, resting-state brain functional connectivity patterns in emerging adolescence that are linked to accelerated maturation and environmental exposures can predict future substance use initiation in youth.

## Data and Code availability

Scripts to generate the results and figures in this study are available at https://github.com/okardan/future_SUI_rsFC. Data tables used in the scripts can be downloaded from the https://nda.nih.gov/study.html?id=2147 after creating an account and being approved on an ABCD data use certificate.

## ABCD Acknowledgements

Data used in the preparation of this article were obtained from the Adolescent Brain Cognitive Development (ABCD) Study (abcdstudy.org), held in the NIMH Data Archive (NDA). This is a multisite, longitudinal study designed to recruit more than 10,000 children age 9 to 10 and follow them over 10 years into early adulthood. The ABCD Study is supported by the National Institutes of Health and additional federal partners under award numbers U01DA041022, U01DA041028, U01DA041048, U01DA041089, U01DA041106, U01DA041117, U01DA041120, U01DA041134, U01DA041148, U01DA041156, U01DA041174, U24DA041123, and U24DA041147. A full list of supporters is available at abcdstudy.org/nih-collaborators. A listing of participating sites and a complete listing of the study investigators can be found at abcdstudy.org/principal-investigators.html. ABCD consortium investigators designed and implemented the study and/or provided data but did not necessarily participate in analysis or writing of this report. This manuscript reflects the views of the authors and may not reflect the opinions or views of the NIH or ABCD consortium investigators.

The ABCD data repository grows and changes over time. The ABCD data used in this report came from NIMH Data Archive Digital Object Identifier 10.15154/8873-zj65.

## Supplementary Material

### 1. Study 1: Constructing the matched control group

The matching was conducted by first generating a pool of all non-SUI participants for each SUI participant who were of the same biological sex, with age difference no more than 6 months, same bracket in household income, same race/ethnicity, and with highest parental education difference no more than 5 years. Income brackets were as follows: 1 = Less than $5,000 ; 2 = $5,000 through $11,999 ; 3 = $12,000 through $15,999 ; 4 = $16,000 through $24,999 ; 5 = $25,000 through $34,999 ; 6 = $35,000 through $49,999 ; 7 = $50,000 through $74,999 ; 8 = $75,000 through $99,999 ; 9 = $100,000 through $199,999 ; 10 = $200,000 and greater. Next, a participant from each of those pools was randomly selected. The process was repeated 10000 times and the list with the greatest number of unique participants constituted the control group used in the study (*N* = 228). The script used for the matching is shared along with the other analysis scripts at https://github.com/okardan/future_SUI_rsFC. There were no differences between the SUI group and the control group in any of the matched categories (age: *p* = .681; sex: *p* = .988; income: *p* = .473; education: *p* = .888; non-Hispanic White: *p* = .508; Black: *p* = 0.849; Hispanic: *p* = .812).

### 2. Study 1: Partial Least Squares analysis

We utilized a partial least squares (PLS) multivariate analysis to distinguish the SUI group from the matched Control group based on their resting state functional brain connectivity in the years prior to SUI. The PLS implementation software was downloaded from Randy McIntosh’s lab at: https://www.rotman-baycrest.on.ca/index.php?section=84. Partial least squares (PLS; [58, 59]) analysis can identify the set of brain functional connections that are maximally related to the group-by-time structure of the data. In PLS, the goal of the analysis is to find weighted patterns of the original variables in the two sets (termed latent variables or LVs) that maximally co-vary with one another. In Task PLS used here (see [59]), these LVs represent a differentiation between levels of experimental design (i. e., two timepoints: baseline and Y2, and two groups: SUI and controls). PLS is computed via singular value decomposition (SVD; [60]) applied to the covariance between the rsFC and the group-by-time contrasts. Applying mean-centering to either groups or timepoints can emphasize the temporal contrast versus the group differences, respectively. In the current analysis, the goal was to allow for a data-driven group-by-time contrast to emerge (i.e. allowing for potential group-by-time interaction). Therefore, only the grand mean was removed from the rsFC values prior to calculation of the covariance matrix X’Y, (where X represents the rsFC matrices and Y represents the group-by-time contrasts). X’Y is then subject to singular value decomposition:

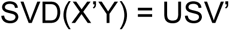

Resulting in a set of orthonormal matrices U and V, as well as a diagonal matrix S of singular values. The number of LVs from the analysis is equal to the smallest rank of its constituent matrices (the rank of the covariance matrix X’Y, which is equal to 4, the degrees of freedom in the experimental design in the current study). The LVs are linear combinations of functional connections across the whole brain whose combinations are differentially instantiated for different groups * time cells. To test the significance of each LV, 1000 covariance matrices were generated by randomly permuting condition labels for the X variables (rsFC). These covariance matrices embody the null hypothesis that there is no relationship between X and Y variables. They were subjected to SVD as before resulting in a null distribution of singular values. The significance of the original LV was assessed with respect to this null distribution. We only interpreted the primary LV in this study, as the other three LVs did not pass the *p* < .01 threshold for significance. The weights in the U singular vector containing the contribution of brain connections to the LV are often referred to as salience.

The reliability with which each functional connection contributes to the overall multivariate pattern (i.e. confidence interval for saliences) was determined with bootstrapping. A set of 1000 bootstrap samples was created by re-sampling subjects with replacement within each group*time cell (i.e. preserving condition labels). Each new covariance matrix was subjected to SVD as before, and the singular vector weights from the resampled data were used to build a sampling distribution of the saliences from the original data set. Saliences that are highly dependent on which participants are included in the analysis will have wide distributions. We then calculated the Bootstrap Ratio Z value by dividing saliences by their estimated standard errors from the bootstraps. The threshold of |Z| > 3 was used for assessing statistical significance of connections as in our prior work [61, 62, 63]. To better visualize the network level contributions to the LV, the proportion of |Z| > 3 connections within each network and also between each pair of networks are presented in Figure 1.

After the PLS, to assess how discriminative the expression of the rsFC pattern is for different types of substances (alcohol, nicotine, cannabis, other drugs, and poly-substance) versus the control group, we calculated a single discriminative expression score for the primary latent variable per participant:

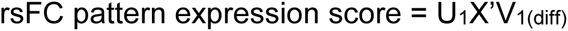

Where X is a 2-by-87153 matrix that contains the connectomes (i.e., flattened rsFC matrices) at baseline and Y2 for the participant, U_1_ (1-by-87153) is the salience vector of the primary LV from Study 1, and V_1(diff)_ (2-by-1) is the difference between SUI and control group’s right-hand singular vectors in the primary LV (see [59]).

### 3. Study 2: Mixed-effects regression analyses

In study 2, we projected the identified rsFC pattern from study 1 onto the fMRI connectivity data in the remainder of participants (i.e., not among the SUI and Control groups) who also had not reported any substance use at baseline or Y2 and had complete data for the environmental and family variables included in the regressions (N = 2,854). This was done again by multiplying the brain salience matrix (U) from the first study (only primary LV) by the rsFC matrix of the study 2 participants at both baseline and Y2 and calculating the weighted sum. The summation weights were the PLS weights from Study 1 corresponding to the SUI group minus the control group so higher projected score corresponds to a Study 2 participant’s connectomes resembling the SUI group more compared to the control group:

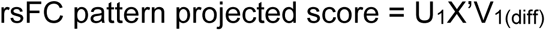

Where X is a 2-by-87153 matrix that contains the connectomes at baseline and Y2 for the participant in Study 2, U_1_ (1-by-87153) is the salience vector of the primary LV from Study 1, and V_1(diff)_ (2-by-1) is the difference between SUI and control group’s right-hand singular vectors in the primary LV from Study 1.

We then regressed these brain scores on the variables of interest (see section 1.3) in two ways. Both analyses involved mixed-effects regressions implemented using lmer function in R package lme4 where standardized beta and p-values were calculated using lmerTest. In both analyses, the model included random intercepts for the data collection site, as well as fixed-effects covariates age, sex, mean frame displacement, household income, and parental education. The difference between the two analyses was whether race/ethnicity was also included as covariates. Race/ethnicity variables are generally included to represent some of the systematic adversities of the marginalized minority groups in the US ([64], see field-wide debates about using race/ethnicity variables as proxies for exposure to unequal levels of adversity, discrimination, and opportunity among marginalized communities in the US [65]). However, because some of the neighborhood variables of interest in the current study including perceived safety, neurotoxins, and ADI may fall within those very adverse experiences, we included the version of the analysis where the variance explained by race/ethnicity was not accounted for.

### 4. Sensitivity analyses: exclusion of family members and equalizing the amount of fMRI data do not change the results

To maximize the sample size and the amount of fMRI data in the main analyses in studies 1 and 2, we utilized all available fMRI runs per participant and included all participants regardless of family nesting due to siblings. We conducted two sensitivity analyses to assess these decisions’ potential impact on the results.

In the first analysis, we equalized the amount of data per participant to one resting-state fMRI run per time-point (i.e., one random run per timepoint was used for participants who had multiple fMRI high quality runs per timepoint). Supplementary Figure S1 shows the PLS primary latent variable in this analysis. Overall, the rsFC pattern was highly correlated with the pattern in the main analysis (r = .80, *p* < .001), and the emerged LV was similar to the main results (i.e., comparing the group by time loadings between Figure 1 and supplementary Figure S1). We then projected this rsFC pattern onto the remainder of the participants and conducted similar regression analyses as in Study 2. The results suggested increased expression of the pattern with higher concentrations of PM2.5, though the other variables in the family, neighborhood pollution, and neighborhood SES domains were not significant (see supplementary Table S1).

In the second sensitivity analysis we randomly retained only one sibling from each family and conducted the PLS (N = 187 SUI and N = 180 control) and the subsequent regression analyses (N = 2,319) on the reduced sample. Again, the primary latent variable in this analysis resembled the main PLS results, with the rsFC pattern highly correlated between the two analyses (r = .73, *p* < .001). The results for this analysis were consistent with the main analysis and are shown in supplementary Figure S2 and Table S2.

**Figure S1.**
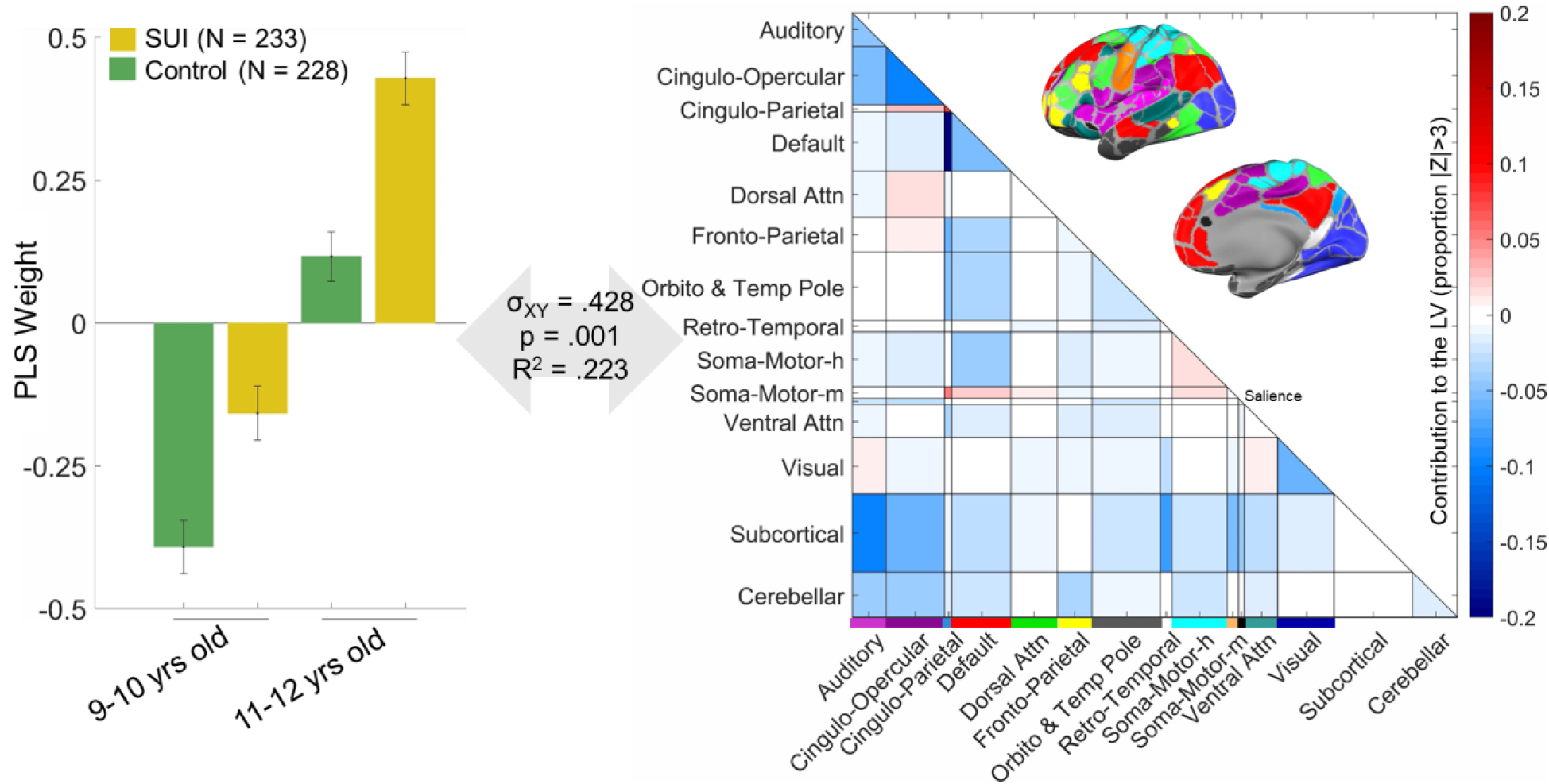
Results of the Partial Least Squares analysis when one run at each time-point for each participant was used to equalize the amount of data used per participant. In the left panel, the group by time PLS weights in the latent variable are shown. The SUI group is in yellow, and the control group is in green; the first two columns show the loadings at 9–10 years of age and the second pair of columns show the loadings at 11-12 years of age. The right panel shows the extent to which the connections across and within brain networks load onto the PLS latent variable. The cortical networks are color-coded, and their anatomical mapping can be seen in the brain surface maps. Networks are defined according to [55]; the ‘unassigned’ set of connections in [55] is labeled here as ‘Orbito and Temp Pole’ as it falls in the orbitofrontal cortex and the temporal pole. The somato-motor networks are labeled with ‘h’ for hand and ‘m’ for mouth. The cross-block covariance is shown as σ_XY,_ and represents the squared singular value for this LV divided by the sum of squared singular values for all LVs. P value is determined by 1000 permutations, and the R^2^ here is the squared Pearson’s correlation (between the left and right-hand sides of the LV (i.e. U and V scores). Z values in the right panel and the error bars in the left panel are estimated using 1000 bootstraps (see methods).

**Table S1.** Results of regressions with the expressed rsFC-SUI pattern from the equal amount of fMRI as the outcome. β is the standardized estimate for the regression coefficient. The difference between Model 1 and 2 is whether race/ethnicity are included as covariates or not. Note: N = 2,854 in both models. Grey highlight shows *p* < .05 within the isolated regression, and addition of * shows *p* < .05 after Holm-Bonferroni correction for all of the regressions.

| Predictor | Model 1<br>*Random effects: Site*<br>*Covariates: Age, Sex, Head motion, Family income, Parental Education* |  |  | Model 2<br>*Random effects: Site*<br>*Covariates: Age, Sex, Head motion, Family income, Parental Education, Race/Ethnicity* |  |  |
| --- | --- | --- | --- | --- | --- | --- |
| | $\beta$ | t | p | $\beta$ | t | p |
| **Family** |  |  |  |  |  |  |
| Parent Hx of Drug Use | 0.011 | 0.701 | 0.483 | 0.020 | 1.285 | 0.199 |
| Parent Hx of Alc Use | 0.002 | 0.139 | 0.891 | 0.002 | 0.152 | 0.879 |
| Family Conflict | 0.009 | 0.581 | 0.562 | 0.011 | 0.732 | 0.464 |
| **Neighborhood (Pollution)** |  |  |  |  |  |  |
| PM2.5 | 0.075 | 3.037 | 0.002* | 0.063 | 2.649 | 0.008 |
| NO2 | 0.038 | 1.580 | 0.114 | 0.020 | 0.859 | 0.390 |
| Lead | 0.037 | 1.638 | 0.101 | 0.018 | 0.813 | 0.416 |
| **Neighborhood (SES)** |  |  |  |  |  |  |
| Area Deprivation | 0.019 | 0.847 | 0.397 | 0.001 | 0.058 | 0.954 |
| Perceived Safety | -0.014 | -0.813 | 0.416 | -0.008 | -0.484 | 0.628 |
| Crime Rate | 0.041 | 1.147 | 0.252 | 0.016 | 0.458 | 0.647 |

**Figure S2.**
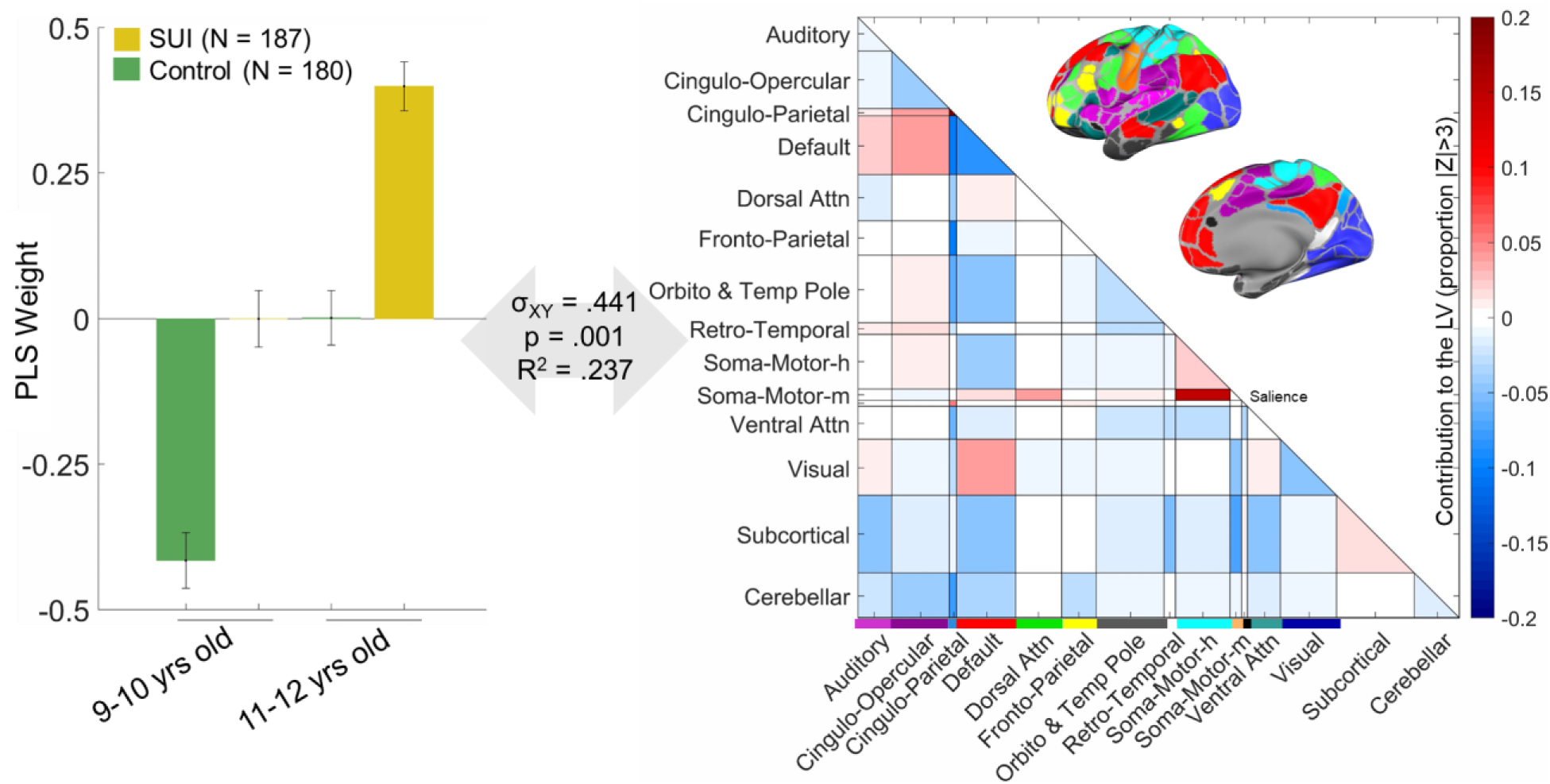
Results of the Partial Least Squares analysis when only one member from each family was used in the analysis. In the left panel, the group by time PLS weights in the latent variable are shown. The SUI group is in yellow, and the control group is in green; the first two columns show the loadings at 9–10 years of age and the second pair of columns show the loadings at 11-12 years of age. The right panel shows the extent to which the connections across and within brain networks load onto the PLS latent variable. The cortical networks are color-coded, and their anatomical mapping can be seen in the brain surface maps. Networks are defined according to [55]; the ‘unassigned’ set of connections in [55] is labeled here as ‘Orbito and Temp Pole’ as it falls in the orbitofrontal cortex and the temporal pole. The somato-motor networks are labeled with ‘h’ for hand and ‘m’ for mouth. The cross-block covariance is shown as σ_XY,_ and represents the squared singular value for this LV divided by the sum of squared singular values for all LVs. P value is determined by 1000 permutations, and the R^2^ here is the squared Pearson’s correlation (between the left and right-hand sides of the LV (i.e. U and V scores). Z values in the right panel and the error bars in the left panel are estimated using 1000 bootstraps (see methods).

**Table S2.**
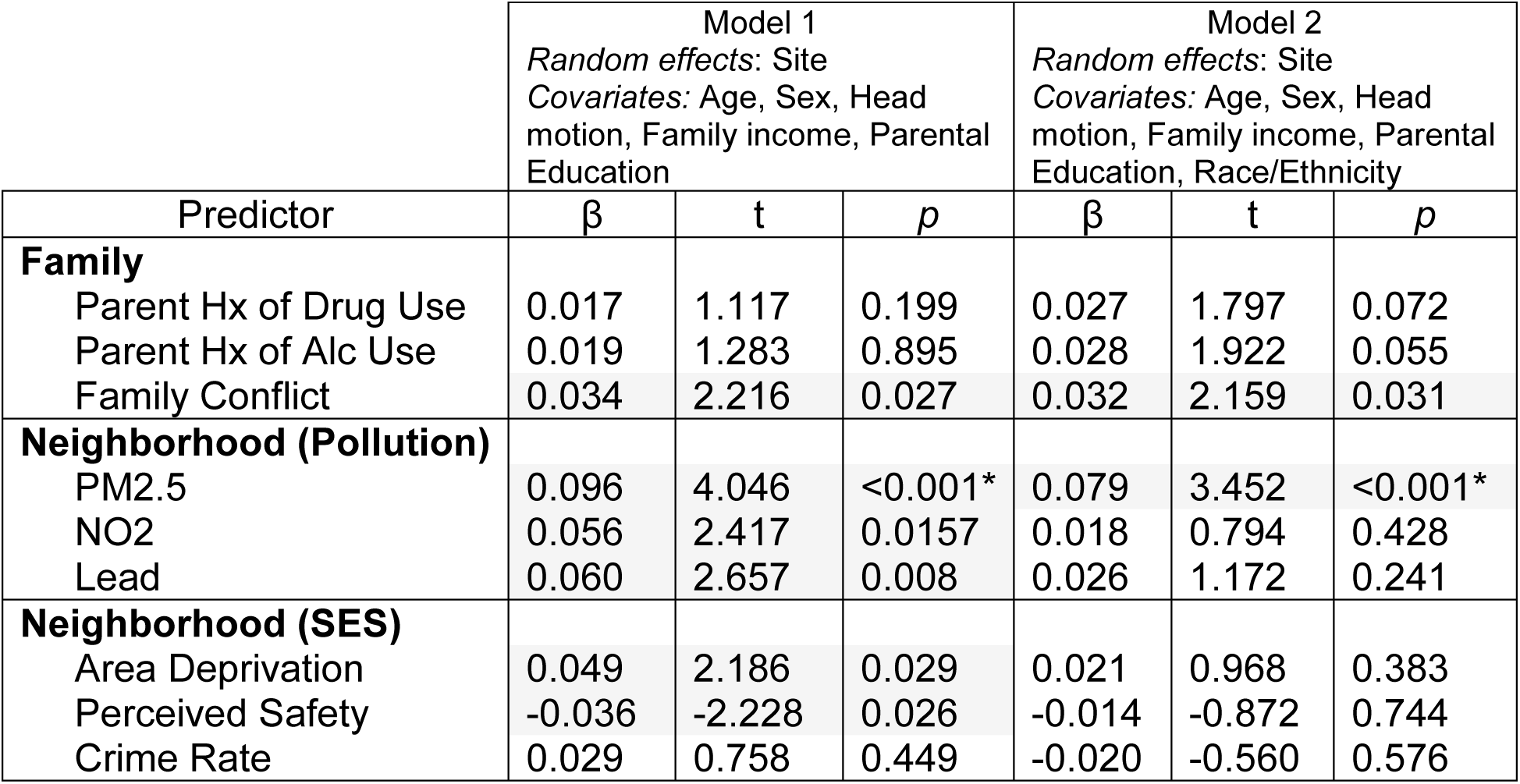
Results of regressions with the expressed rsFC-SUI pattern from the one-family-member analysis as the outcome. β is the standardized estimate for the regression coefficient. The difference between Model 1 and 2 is whether race/ethnicity are included as covariates or not. Note: N = 2,319 in both models (due to exclusion of siblings). Grey highlight shows *p* < .05 within the isolated regression, and addition of * shows *p* < .05 after Holm-Bonferroni correction for all of the regressions.

